# Violent victimisation in post-traumatic stress disorder: a Swedish register-based cohort study

**DOI:** 10.64898/2026.10.29.26366725

**Authors:** Anabelle Paulino, Amir Sariaslan, Ralf Kuja-Halkola, Brian M. D’Onofrio, Zheng Chang, Ebba du Rietz, Paul Lichtenstein, Isabell Brikell, Seena Fazel

## Abstract

**Background:** Post-traumatic stress disorder (PTSD) is associated with various adverse outcomes, but links with subsequent violent victimisation are uncertain.

**Methods:** We linked data from Swedish sociodemographic and medical registers to create a matched cohort of people with diagnosed PTSD and a comparison sample matched on birth year, sex, and county of residence in the year preceding diagnosis (or matching in the case of undiagnosed individuals). We calculated the cumulative incidence of violent victimisation (defined using ICD codes registered at hospitalisation) and conducted stratified Cox regression analyses, adjusted for parental factors, psychiatric comorbidities, and history of violent victimisation.

**Findings:** The cohort contained 29,688 individuals diagnosed with PTSD, who were matched with 296,880 individuals without PTSD (78.6% female), and followed for up to 34 years. Individuals with PTSD had higher cumulative incidences of violent victimisation after their index diagnosis (18.7% compared to 2.7% at 25 years follow-up). Individuals with both a diagnosis of PTSD and history of violent victimisation had the highest cumulative incidence (28.7%). After adjustment for parental characteristics, prior psychiatric comorbidity and prior violent victimisation, PTSD was associated with an adjusted hazard ratio of 4.0 (95% CI 3.7–4.4).

**Interpretation:** Preventing subsequent violent victimisation among individuals diagnosed with PTSD should be considered by clinical services and incorporated into violence prevention strategies.

**Funding:** AP is funded by the Wellcome Trust (grant code 218497/Z/19/Z). SF and AS are supported by the NIHR Oxford Health Biomedical Research Centre (grant BRC-1215-20005). No funder(s) listed had any role in the design or implementation of this study.

## Introduction

Post-traumatic stress disorder (PTSD) is a public health issue associated with substantial societal costs and personal suffering for those affected.^1^ Individuals with PTSD have a higher risk of adverse outcomes such as psychiatric comorbidities and complications arising from physical health problems, as well as substance use disorders, self-harm, suicide, and violence perpetration.^2–6^ However, an outcome of public health importance not widely studied in relation to PTSD is violent victimisation.^6^

Violent victimisation is associated with subsequent diagnoses of common psychiatric disorders (including PTSD), suicidality, and premature mortality.^7^ Conversely, individuals with mental health disorders have reported increased risks of subsequent violent victimisation, highlighting the bidirectional associations of risk.^8,9^ Specific mechanisms underlying victimisation, including revictimisation, in individuals include numbing from dissociation, emotional numbing, and disorientation.^10^ Revictimisation can further contribute to future victimisation risks, especially in people with PTSD.^10,11^ Experimental studies have shown that individuals with PTSD display heightened sensitivity towards potential threats and difficulties in disengaging from threatening trauma-related stimuli,^2,12^ which may explain potential links between PTSD and revictimisation.

Current evidence on the association between PTSD and subsequent violent victimisation is limited by small or selected sample sizes.^13–15^ Another limitation relates to the timing of the victimization, which is unclear in cross-sectional studies or those reporting lifetime violent victimisation. A Danish population study found links between a broad category of anxiety-related disorders and victimisation^16^ in a longitudinal design but did not report PTSD as a separate diagnostic category.

Given the association of violent victimisation with subsequent diagnoses of psychiatric disorders, including PTSD,^8,17^ the association between PTSD and violent victimisation can be considered as part of a cycle of abuse and violence. Investigating the association between PTSD and violent victimisation can therefore inform treatment for individuals at different points of PTSD diagnosis and violent victimisation exposure, including preventing revictimisation.^18^ Furthermore, given differential risks of other violence-related outcomes by sex, comorbid substance use disorder, and history of violence, we sought to ascertain whether subsequent violent victimisation may also vary within these subgroups.^5^ Thus, to investigate the association between a PTSD diagnosis and subsequent violent victimisation, we conducted a register-based cohort study using nationwide Swedish data.

## Methods

### Data sources

We used national Swedish registers containing person-specific sociodemographic and medical information, linked via a pseudonymised version of the individual personal identification number assigned at birth or upon immigration into Sweden.^19^

We obtained date of birth, legal sex (hereto after referred to as “sex”), and birth country for individuals living in Sweden from 1947 onwards through the Total Population Register (TPR).^20^ The TPR has effectively 100% coverage for births in Sweden and 95% coverage for immigration into Sweden, with this proportion increasing over time.^20^ We connected individuals born from 1932 onwards and registered as living in Sweden from 1961 onwards to their family members through the Multi-Generation Register (MGR).^21^ MGR contains information on 97% of mothers and 95% of fathers of the index individuals.^21^

We retrieved information on psychiatric inpatient hospitalisations from 1973 onwards and outpatient visits from 2001 onwards from the National Patient Register (NPR).^22^ Inpatient coverage for psychiatric disorders began at around 86% at inception of the register and increased to 100% by 1987, while outpatient coverage from public caregivers is nearly 100%.^22^ Psychiatric diagnoses in the NPR are listed using the ICD format and include diagnoses from the eighth (ICD-8), ninth (ICD-9), and tenth (ICD-10) Swedish editions. The Cause of Death register, which contains information on deaths in Sweden since 1952, was used to obtain information on date of death.^23^ As of 2012, this register lists all individuals who have died in Sweden regardless of residency.^23^

For information on family income and education by calendar year, we used the Integrated Database for Health Insurance and Labour Market Studies (LISA).^24^ Since LISA only contains information from 1990 onwards,^24^ data on income, education, and employment pre-1990 was obtained through other relevant databases from Statistics Sweden. Information from LISA covers all individuals aged 16 and older from 1990 onwards, in addition to those aged 15 and older from 2010 onwards.^24^ Information on county of residence was obtained from Demografiska Statistikområden (DeSO).^25^ The Migration Register, also managed by Statistics Sweden, contains information on immigration and emigration, including date, type, and location, from around 1968 onwards.^26^

We obtained access to this pseudonymised data after approval from the Swedish Ethical Review Authority (Dnr 2020–06540; Dnr 2022-06204-02). In accordance with Swedish law, informed consent is waived for register-based research using pseudoanonymised data.^27^

### Study population

We derived the study population by selecting individuals born in Sweden from 1 January 1973 until 31 December 2005. We chose this date range in line with the PTSD diagnosis (included from 1987 onwards) being included in ICD-9 and ICD-10, while also allowing for sufficient follow-up time to assess the violent victimisation outcome. We then excluded adopted individuals and those with information missing on one or both parents, in order to be able to ascertain biological parental and sibling relationships.

Our analytic cohort was created by selecting all individuals with a primary or secondary PTSD diagnosis as recorded in the NPR. PTSD diagnoses in the NPR have been validated for epidemiological research, with a positive predictive value of 84% (95% confidence interval (CI) 79%–90%).^28^ and are recorded under ICD-9 code 309B or ICD-10 code F43.1. For individuals with multiple PTSD diagnoses, the first recorded PTSD diagnosis was used. We excluded PTSD diagnoses before age 10, as childhood PTSD diagnoses are more rare and could have been misdiagnoses.^29^ We then matched without replacement each individual diagnosed with PTSD to ten individuals from the general population who were alive, residing in Sweden, and had not been diagnosed with PTSD at the time of matching. Individuals were matched on sex, birth year, and county of residence in one of 21 counties in the year preceding diagnosis of the index person.

See **Appendix Figure S1** for a visual representation of the cohort derivation.

### Measures

Our main outcome was violent victimisation after index PTSD diagnosis. We defined violent victimisation as an inpatient or outpatient visit recorded in the NPR relating to an externally and deliberately inflicted injury as per previous work.^8^ To differentiate violent victimisation outcomes in ICD classification system as being independent events from PTSD diagnoses, we only counted violent victimisation outcomes at least one month after PTSD diagnoses. We used codes E960–E969 (ICD-8 and ICD-9), X85–X99, and Y00–Y09 (ICD-10) for these outcomes.

Covariates used in these analyses were parental characteristics, psychiatric comorbidities, and history of violent victimisation. Within parental characteristics included in analyses, these were low family income, low parental education, and parental immigration status, as obtained from LISA and census data and consistent with previous literature definitions.^8^ Individuals were considered children of immigrants if at least one parent was born outside of a Nordic country. Low parental education was defined as at least one parent completing less than secondary school education, or if at least one parent had information on parental education missing. Low parental income was defined by the family income measurement being in the bottom decile.

Psychiatric comorbidities included were substance use disorder (SUD), major depressive disorder (MDD), bipolar disorder, schizophrenia, attention deficit hyperactivity disorder (ADHD)/conduct disorder, and anxiety disorders as recorded through inpatient or outpatient visits in the NPR. (Other neurodevelopmental disorders were considered outside the scope of the study.) Once individuals were diagnosed, we considered these diagnoses as lifetime. These diagnoses were not considered hierarchical or mutually exclusive, meaning that individuals could have multiple diagnoses, each of which would count towards the respective diagnostic group. Only diagnoses that preceded PTSD (or matching, in the case of undiagnosed individuals) were included in analyses to help establish the temporality of these specific covariates relative to exposure. We defined history of violent victimisation as any diagnosis of violent victimisation occurring at least a month before the date of the index person’s PTSD diagnosis. See **Appendix Table S1** for the specific ICD-8, ICD-9, and ICD-10 codes used.

### Statistical analyses

We followed individuals in the matched cohort from the date of PTSD diagnosis for the index person until the first of the following: violent victimisation, death, emigration, or 31 December 2020 (end of follow-up period), or in the instance of control individuals, future PTSD diagnosis. We then generated Kaplan-Meier cumulative incidence curves of violent victimisation, first for individuals with and without PTSD diagnoses. We also did stratified analyses of individuals with and without histories of violent victimisation. Next, we implemented stratified Cox regression with the underlying time scale of time since diagnosis, making comparisons within strata which allowed for baseline hazard rates to vary between them. Each set of one individual diagnosed with PTSD and their ten corresponding matched individuals comprised a stratum. The outputs of stratified Cox regressions were hazard ratios (HRs) with 95% CIs.

The minimally-adjusted matched cohort model implicitly adjusted for birth year, sex, and county of residence in the year preceding diagnosis through matching. Adjustments were sequentially added in the following order: parental characteristics, psychiatric comorbidities diagnosed prior to PTSD or matching, and history of violent victimisation.

We then ran various stratified analyses on the fully-adjusted model to investigate potential differences in the association between PTSD diagnosis and violent victimisation in subgroups. We ran stratified analyses by sex, SUD diagnosis prior to PTSD diagnosis, and history of violent victimisation.

Sensitivity analyses on the matched cohort included birth cohort restrictions from 1983 onwards, to limit the definition of PTSD to ICD-10 and to account for potential birth cohort effects from older birth cohorts, and analyses on only individuals with a more stringent PTSD diagnosis (represented as the primary diagnosis in the NPR).

We performed data linkage and analyses using SAS version 9.4, and the “survival” and “survminer” packages in R version 4.3.1.^30–32^

## Results

There were 326,568 total individuals included in the matched cohort. Among them, 29,688 were diagnosed with PTSD and 296,880 were matched individuals without PTSD diagnoses at start of follow-up. Follow up was for a median 4.3 (IQR 6.2, range 0-34) years. Overall, 4,748 individuals experienced violent victimisation during follow-up. In those with PTSD diagnoses, 7.1% experienced violent victimisation during follow-up, while in the age, sex, and county of residence-matched control group, 0.9% individuals experienced violent victimisation (**Table 1**; information on lifetime psychiatric comorbidities in **Appendix Table S2)**.

**Table 1:** Sample characteristics of people with diagnoses of PTSD and a matched cohort in the general population.

|  | **PTSD *(% total)*** | **No PTSD *(% total)*** |
|---|---|---|
| **TOTAL** | 29,688 *(9.1%)* | 296,880 *(90.9%)* |
| Median [IQR] diagnosis age, years | 25.6 [12.2] | .. |
| Median (IQR) [range] follow-up time, years | 4.0 (6.0) [0.0 - 33.9] | 4.4 (6.3) [0.0 - 33.9] |
| Total follow-up time, person-years | 156,517 | 1,665,075 |
|  | **PTSD *(% PTSD)*** | **No PTSD *(% No PTSD)*** |
| **Sex** |  |  |
| Male | 6,358 *(21.4%)* | 63,580 *(21.4%)* |
| Female | 23,330 *(78.6%)* | 233,300 *(78.6%)* |
| **Parental characteristics** |  |  |
| Parental immigration | 4,898 *(16.5%)* | 37,123 *(12.5%)* |
| Low parental education | 2,259 *(7.6%)* | 15,158 *(5.1%)* |
| Low family income | 3,352 *(11.3%)* | 21,601 *(7.3%)* |
| Missing information | 72 *(0.2%)* | 447 *(0.2%)* |
| **Psychiatric comorbidities prior to matching** |  |  |
| Substance use disorder | 7,125 *(24.0%)* | 11,562 *(3.9%)* |
| Schizophrenia | 169 *(0.6%)* | 299 *(0.1%)* |
| Bipolar disorders | 2,263 *(7.6%)* | 2,713 *(0.9%)* |
| Major depressive disorder | 13,808 *(46.5%)* | 18,845 *(6.3%)* |
| ADHD/conduct disorders | 5,384 *(18.1%)* | 10,193 *(3.4%)* |
| Anxiety disorders | 15,373 *(51.8%)* | 22,665 *(7.6%)* |
| **Violent victimisation** |  |  |
| Prior violent victimisation | 4,065 *(13.7%)* | 6,143 *(2.1%)* |
| Violent victimisation during follow-up | 2,094 *(7.1%)* | 2,654 *(0.9%)* |
| Median [IQR] age at victimisation during follow-up in years | 25.6 [11.6] | 24.8 [9.7] |

IQR: interquartile range; SD: standard deviation; ADHD: attention deficit hyperactivity disorder.

Individuals with PTSD had higher cumulative incidences of violent victimisation during follow-up. The cumulative incidence of violent victimisation at 5 years in individuals with a PTSD diagnosis was 7.5% (95% CI 7.2%–7.9%) compared to 0.9% (95% CI 0.9%–0.9%) in individuals without PTSD. This rose to 18.7% (95% CI 16.0%–21.3%) in individuals with PTSD compared to 2.7% (95% CI 2.4%–3.1%) in individuals without PTSD at 25 years of follow-up (see **Figure 1** and **Appendix Table S3**).

**Figure 1:**
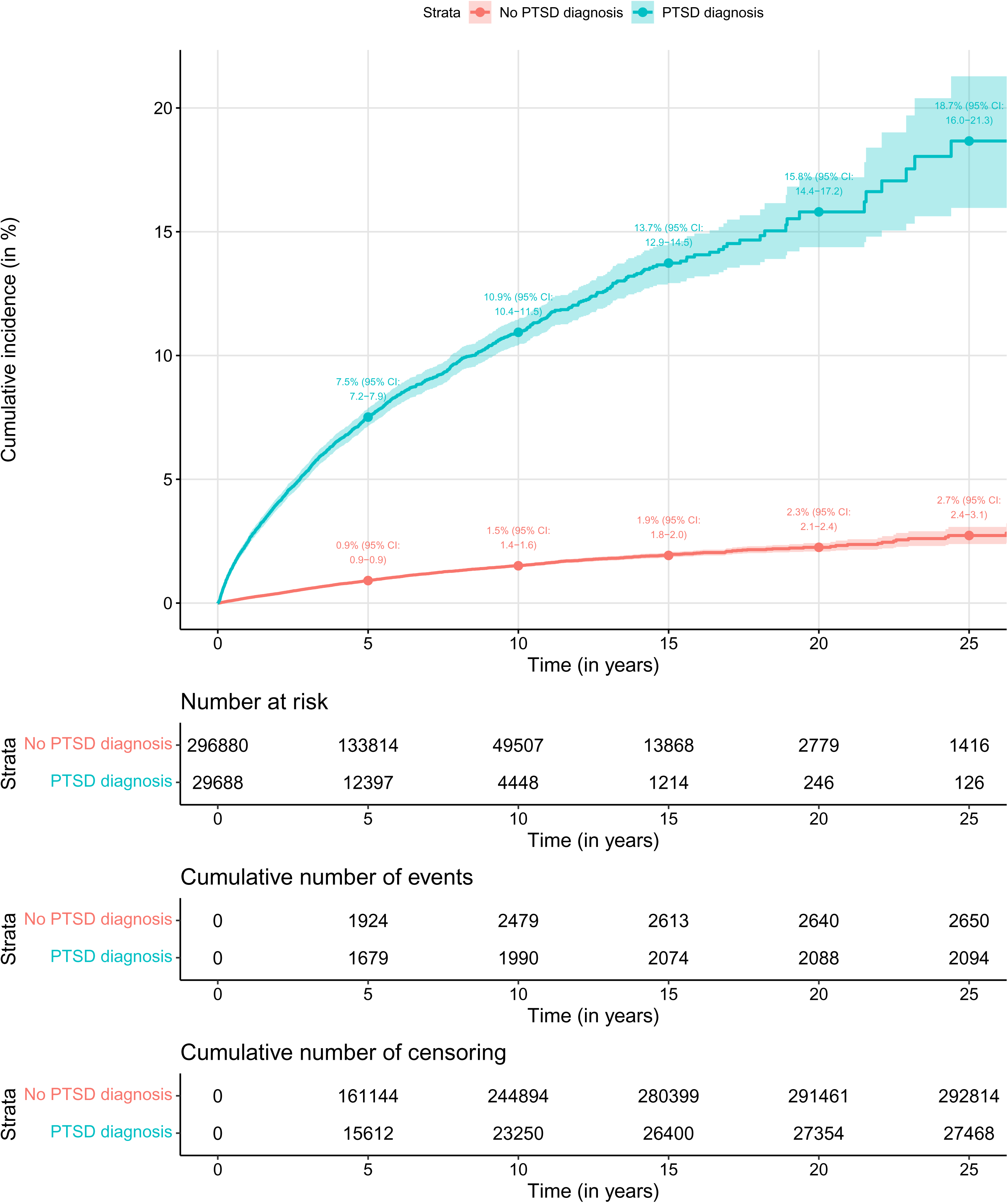
Cumulative incidence of violent victimisation by PTSD diagnosis Although individuals were followed for up to approximately 34 years, we used a 25-year time point cut-off for the cumulative incidence plot due to no more change in cumulative incidence among individuals with PTSD past the 25-year time point.

Individuals with both PTSD diagnoses and history of violent victimisation had the highest cumulative incidence of violent victimisation (28.7%, 95% CI 24.4%–32.8% at 25 years of follow-up) (**Figure 2**; **Appendix Table S4**). Lower cumulative incidences of violent victimisation were found in both individuals with PTSD diagnosis and no history of violent victimisation (16.9%, 95% CI 14.1%–19.6% at 25 years of follow-up) and individuals with a history of violent victimisation but no PTSD diagnosis (9.3%, 95% CI 7.6%–11.0%). However, both these groups still had elevated risks throughout the entire follow-up period when compared to individuals who did not have either a PTSD diagnosis or history of violent victimisation (2.6%, 95% CI 2.2%–3.0%).

**Figure 2:**
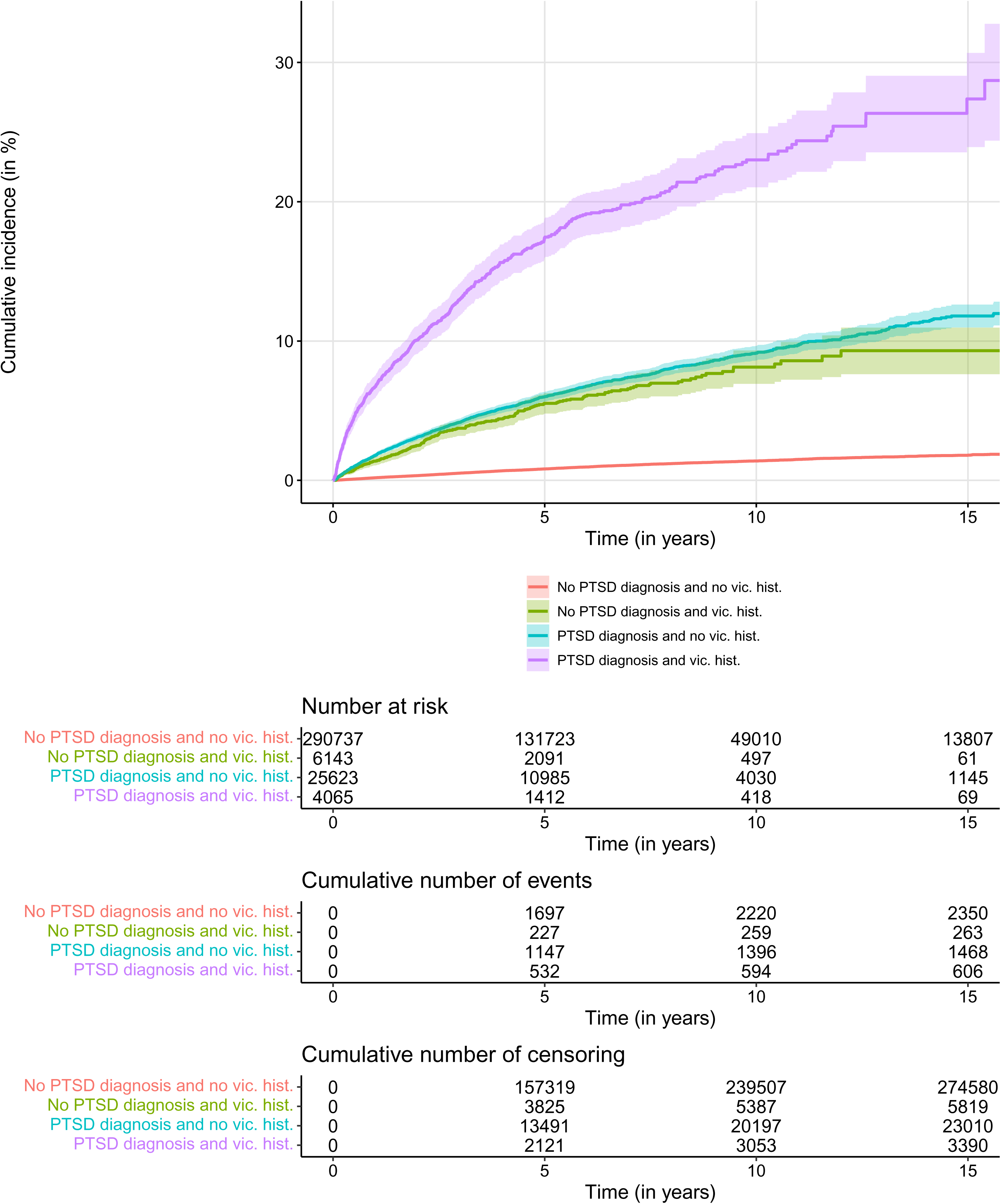
Cumulative incidence of violent victimisation by PTSD diagnosis and history of violent victimisation We used a 15-year cut-off point due to Swedish data privacy laws surrounding the reporting of few Ns.

Individuals with PTSD had consistently higher risks of violent victimisation compared to undiagnosed matched individuals, including after adjusting for parental characteristics, psychiatric comorbidities prior to PTSD diagnosis, and history of violent victimisation (HR 4.0 (95% CI 3.7–4.4)) (**Table 2**) In stratified analyses, females with PTSD had a higher hazard ratio of violent victimisation compared with females without PTSD (HR 4.7 (95% CI 4.3–5.2) than the corresponding comparison among males (HR 2.7 (95% CI 2.3–3.1)). Similarly, the HRs of violent victimisation associated with PTSD were higher among individuals with no prior SUD and no history of violent victimisation than their counterparts with prior SUD and history of violent victimisation respectively (**Table 3**). Both the restricted birth cohort of individuals born 1983 or later (**Appendix Table S5**) and alternative (primary diagnosis) definition of PTSD (**Appendix Table S6**) showed similar results to the original matched cohort analyses.

**Table 2:** Hazard ratios for the association of PTSD and subsequent violent victimisation.

|  | **Cohort of PTSD and matched general population** |  |  |
|---|---|---|---|
| **Statistical adjustment** | N PTSD | N TOTAL | HR (95% CI) |
| Basic model | 29,688 | 326,568 | 8.4 (7.9 – 8.9) |
| Parental characteristics included | 29,616 | 326,049 | 8.1 (7.7 – 8.6) |
| Parental characteristics and psychiatric comorbidities prior to matching included | 29,616 | 326,049 | 4.6 (4.2 – 4.9) |
| Parental characteristics, psychiatric comorbidities prior to matching, and history of violent victimisation included | 29,616 | 326,049 | 4.0 (3.7 – 4.4) |

Basic model (matched cohort): implicitly adjusted for age, sex, and county of residence in the year of diagnosis through matching.

**Table 3:** Stratified analyses on fully-adjusted matched cohort models investigating links between PTSD diagnosis and subsequent violent victimisation.

|  | **Cohort of PTSD and matched general population** |  |  |
|---|---|---|---|
|  | N PTSD | N TOTAL | HR (95% CI) |
| **Model 5** (stratified by sex) |  |  |  |
| *Male* | 6,332 | 69,804 | 2.7 (2.3 – 3.1) |
| *Female* | 23,284 | 256,245 | 4.7 (4.3 – 5.2) |
| **Model 6** (stratified by prior SUD) |  |  |  |
| *No prior SUD* | 22,514 | 307,403 | 4.8 (4.4 – 5.2) |
| *Prior SUD* | 7,102 | 18,646 | 2.1 (1.8 – 2.5) |
| **Model 7** (stratified by violent victimisation history) |  |  |  |
| *No prior violent victimisation* | 25,564 | 315,873 | 4.3 (4.0 – 4.7) |
| *Prior violent victimisation* | 4,052 | 10,176 | 2.5 (2.0 – 3.1) |

Fully-adjusted (matched cohort): implicitly adjusted for age, sex, and county of residence in the year of diagnosis through matching, and adjusted for parental characteristics, psychiatric comorbidities prior to matching, and history of violent victimisation

## Discussion

In this cohort study of 326,568 individuals in the Swedish population, we investigated the association between PTSD and subsequent violent victimisation over up to 34 years of follow-up. We found that individuals with PTSD were at increased risk for subsequent violent victimisation compared with individuals matched for sex and age without a PTSD diagnosis in the general population. The 5-year cumulative incidence of violent victimisation in individuals with a PTSD diagnosis was 7.5%, and the hazards of violent victimisation were elevated around fourfold compared with the general population after adjustment for background social and psychiatric factors. These absolute and relative risks indicate that subsequent violent victimisation is a clinically relevant outcome, and risks are at least as high as other adverse outcomes reported in people with PTSD. For example, other work has reported comparable cumulative incidences for violent perpetration, and relative risks are in a comparable range with violent perpetration, suicide, and self-harm.^4,5^

We found that, in stratified analyses, women with PTSD were at greater relative risk of violent victimisation when compared to their male counterparts. This is broadly consistent with a meta-analysis showing elevated risks of interpersonal violent victimisation in women with PTSD and other research on risk of revictimisation stratified by gender.^11,13^ Stratified analyses furthermore showed that, in individuals with comorbid SUD or history of violent victimisation, a PTSD diagnosis itself did not significant alter the association between PTSD and violent victimisation. This could be explained by victimisation history or comorbid SUD having a stronger link with subsequent violent victimisation than a PTSD diagnosis. In relation to violent victimisation, individuals with both PTSD diagnoses and prior violent victimisation had highest absolute risks of subsequent violent victimisation. With other work additionally showing associations between violent victimisation and subsequent PTSD, the current findings support a model of a cycle of trauma, PTSD, and victimisation in individuals who are repeat victims of violence.

There are several implications for clinical practice and public health. In individuals who develop PTSD symptoms and a diagnosis, clinical services should consider risks of violent victimisation. Some treatments exist that can mitigate this risk,^33^ but their wider adoption will require further trials. Furthermore, the study findings support public mental health measures that could reduce incident PTSD itself.^1^ These measures might include increasing resilience, and other psychological and pharmacological treatments in people exposed to violence.^2,34^ Finally, there can be a role of prognostic models, which in the context of PTSD can provide a framework for risk assessment and support for clinical decision-making.^35^ Within risk assessment and treatment in individuals with a PTSD diagnosis, the results of this study indicate that particular subgroups to prioritise can include individuals with history of violent victimisation, as well as women.

Strengths of this study include the use of longitudinal data, a large representative population, random selection, standardised definitions, and adjustment for parental characteristics. However, one limitation of the study relates to country of birth or residence of individuals included. We were only able to include individuals born in Sweden with information on their biological parents due to the study of parental factors as a covariate. Because immigrants to Sweden are not included, this study therefore underrepresents individuals whose PTSD may have resulted from potential adverse experiences associated with migration such as exposure to war, severe hardship, or refugee status. Given that migrants, and particularly refugees, have shown elevated risks of healthcare utilisation relating to PTSD (ranging from approximately three-fold for non-refugee migrants to 8-fold risk in refugee migrants compared to individuals born in Sweden), this population is particularly important to investigate.^36^ The inclusion of only individuals born in Sweden could additionally affect the generalisability of findings to other populations, as the sample of individuals with PTSD diagnoses in Sweden may not be representative of PTSD-diagnosed individuals elsewhere due to the low exposure to combat and war within Sweden. A related limitation to this is the lack of information on ethnicity in our study, as this information is not collected in Swedish registers.^37^ While variables including country of birth or residence could be considered rough proxies for race or ethnicity in population research, these may not fully capture the nuance of ethnicity in the link between PTSD and violent victimisation.^37^

Reverse causality is another potential limitation, which we addressed using a period of one month after PTSD diagnosis for measurement of the outcome. Further, with stratified analyses by history of violent victimisation, we demonstrated that even in the absence of violent victimisation history, there remains an association between PTSD and violent victimisation.

We were not able to determine the underlying causes of PTSD or violent victimisation. This could have indicated whether the PTSD resulted from a traumatic event involving violent victimisation, and stratify risks by exposure event. As recorded in the patient register, PTSD or violent victimisation had to be severe enough to lead to specialist or inpatient medical treatment for inclusion, meaning our definitions would have captured the more severe end of the spectrum. Nevertheless, these diagnoses are more relevant from a service-perspective as they involve contact with healthcare, and also provide opportunities for intervention. Furthermore, in terms of violent victimisation, evidence has shown high genetic correlation between the register-based definition and broader measures of police-reported violent victimisation in Finland, which therefore provides some validation to the measure used in this study^38^. The timing of violent victimisation outcomes are dependent on when medical treatment was sought, so timing of these diagnoses may be delayed. The underreporting of violent victimisation, both at an individual and population level, is also a limitation. However, this underreporting in people would likely be similar in people with and without PTSD diagnosis, and thus unlikely to materially affect relative risk estimates.

In conclusion, we have shown that there is an association between PTSD and subsequent violent victimisation using a longitudinal design. This association remained after accounting for parental factors, psychiatric comorbidities, and history of violent victimisation. Further research is warranted to further disentangle the contribution of these factors, and whether and what treatments can mitigate risk of violent victimisation after PTSD.

## Data Availability

Individual-level data is not publicly available due to Swedish privacy laws. Interested researchers can apply for data access through Statistics Sweden (https://www.scb.se/en/services/ordering-data-and-statistics/microdata/) and the Swedish National Board for Health and Welfare (https://www.socialstyrelsen.se/en/statistics-and-data/registers/).

## Contributors

SF and AP were responsible for the conception of the study. Formal analysis was done by AP, who designed the study with SF, AS, RKH, PL and IB. AP and AS had access to the data; AP accessed it and verified it with AS. AP drafted the paper, and all authors reviewed, revised, and approved the final manuscript. SF provided overall supervision. All authors were responsible for the decision to submit for publication.

## Acknowledgements

AP is on a PhD studentship funded by the Wellcome Trust (grant code 218497/Z/19/Z). For the purpose of open access, the author has applied a CC BY public copyright licence to any Author Accepted Manuscript version arising from this submission. SF and AS are supported by the NIHR Oxford Health Biomedical Research Centre.

The funder(s) had no role in the design or implementation of this study.

