## Appendix for "Violent victimisation in post-traumatic stress disorder: a Swedish register-based cohort study"

**Table S1:** ICD codes for diagnoses used in analyses

|  | **ICD-8 (1981–1986)** | **ICD-9 (1987–1996)** | **ICD-10 (1997–2020)** |
| --- | --- | --- | --- |
| *PTSD* | N/A | 309B | F43.1 |
| *SUD* | 303 and 304 | 303, 304, 305A, and 305X | F10–F19 (excluding F17) |
| *Depression* | 296.2 and 300.4 | 296B, 300E, and 311 | F32–F39 (excluding F32.3) |
| *Bipolar disorder* | 296 (excluding 296.2) | 296 (excluding 296B) | F30–F31 |
| *Schizophrenia* | 295 | 295 | F20 |
| *ADHD/conduct disorder* | N/A | 312 and 314 | F90–F91 |
| *Anxiety disorders* | 300 (excluding 300.4) | 300 (excluding 300E) | F40–F42, F44–F45, F48 |
| *Violent victimisation** | E960–E969 | E960–E969 | X85–X99 and Y00–Y09 |

*Violent victimisation was represented using a composite flag variable (F57) in the NPR flag register. These ICD codes are the ones that fall under this composite variable, although we did not have access to the individual ICD diagnoses themselves in the register data provided.

**Table S2:** Lifetime prevalence of psychiatric comorbidities vs. PTSD diagnosis and violent victimisation

|  | **Matched cohort (N = 326,568)** | |
| --- | --- | --- |
|  | **PTSD (% PTSD)**  **n = 29,688** | **No PTSD (% No PTSD)**  **n = 296,880** |
| Schizophrenia | 380 *(1.3%)* | 452 *(0.2%)* |
| Bipolar disorders | 4,324 *(14.6%)* | 4,854 *(1.6%)* |
| Major depressive disorder | 20,380 *(68.6%)* | 29,745 *(10.0%)* |
| Anxiety disorders | 22,278 *(75.0%)* | 36,247 *(12.2%)* |
| ADHD/conduct disorders | 9,388 *(31.6%)* | 15,591 *(5.3%)* |
| Substance use disorder | 10,128 *(34.1%)* | 16,597 *(5.6%)* |
|  | **Violent victimisation (% victimised)**  **n = 4 748** | **No violent victimisation (% not victimised)**  **n = 321 820** |
| Schizophrenia | 51 *(1.0%)* | 781 *(0.2%)* |
| Bipolar disorders | 529 *(11.1%)* | 8,643 *(2.7%)* |
| Major depressive disorder | 2,268 *(47.8%)* | 47,587 *(14.8%)* |
| Anxiety disorders | 2,682 *(56.5%)* | 55,843 *(17.4%)* |
| ADHD/conduct disorders | 1,538 *(32.4%)* | 23,441 *(7.3%)* |
| Substance use disorder | 2,249 *(47.4%)* | 24,476 *(7.6%)* |

**Table S3:** Cumulative incidence of violent victimisation, by PTSD diagnosis

|  | Cumulative incidence (95% CI) | |
| --- | --- | --- |
| Time since follow-up | PTSD diagnosis | No PTSD diagnosis |
| 1 year | 2.5% (2.3% - 2.6%) | 0.2% (0.2% - 0.2%) |
| 2.5 years | 4.7% (4.5% - 5.0%) | 0.5% (0.5% - 0.5%) |
| 5 years | 7.5% (7.2% - 7.9%) | 0.9% (0.9% - 0.9%) |
| 10 years | 10.9% (10.4% - 11.5%) | 1.5% (1.4% - 1.6%) |
| 15 years | 13.7% (12.9% - 14.5%) | 1.9% (1.8% - 2.0%) |
| 20 years | 15.8% (14.4% - 17.2%) | 2.2% (2.1% - 2.4%) |
| 25 years | 18.7% (16.0% - 21.3%) | 2.7% (2.4% - 3.1%) |

**Table S4:** Cumulative incidence of violent victimisation, by PTSD diagnosis and history of violent victimisation

|  |  | Cumulative incidence (95% CI) | |
| --- | --- | --- | --- |
| Time since follow-up | History of violent victimisation | PTSD diagnosis | No PTSD diagnosis |
| 1 year | *Yes* | 6.7% (6.0% - 7.5%) | 1.4% (1.1% - 1.7%) |
|  | *No* | 1.8% (1.6% - 2.0%) | 0.2% (0.2% - 0.2%) |
| 2.5 years | *Yes* | 11.4% (10.4% - 12.5%) | 3.3% (2.8% - 3.8%) |
|  | *No* | 3.6% (3.4% - 3.9%) | 0.4% (0.4% - 0.5%) |
| 5 years | *Yes* | 17.4% (16.0% - 18.8%) | 5.5% (4.8% - 6.3%) |
|  | *No* | 6.0% (5.7% - 6.3%) | 0.8% (0.8% - 0.9%) |
| 10 years | *Yes* | 23.0% (21.0% - 24.9%) | 8.1% (7.0% - 9.3%) |
|  | *No* | 9.1% (8.6% - 9.7%) | 1.4% (1.3% - 1.5%) |
| 15 years | *Yes* | 27.4% (23.9% - 30.7%) | 9.3% (7.6% - 11.0%) |
|  | *No* | 11.8% (11.0% - 12.6%) | 1.8% (1.7% - 1.9%) |
| 20 years | *Yes* | 28.7% (24.4% - 32.8%) | 9.3% (7.6% - 11.0%) |
|  | *No* | 13.9% (12.4% - 15.3%) | 2.1% (2.0% - 2.2%) |
| 25 years | *Yes* | 28.7% (24.4% - 32.8%) | 9.3% (7.6% - 11.0%) |
|  | *No* | 16.9% (14.1% - 19.6%) | 2.6% (2.2% - 3.0%) |

**Table S5-S6: Sensitivity analyses**

**Table S5:** Birth cohort (1983-)

| **Model specification** | **Matched Cohort**  **(N = 240,999)** |
| --- | --- |
|  | HR (95% CI) |
| Minimally-adjusted | **7.9** (7.4 - 8.5) |
| Parental characteristics | **7.6** (7.1 - 8.1) |
| Parental characteristics + psychiatric comorbidities | **4.5** (4.2 - 5.0) |
| Parental characteristics + psychiatric comorbidities + history of violent victimization (fully-adjusted) | **4.0** (3.7 - 4.4) |
| Fully-adjusted (stratified by sex) |  |
| *Male* | **2.6** (2.2 - 3.0) |
| *Female* | **4.8** (4.3 - 5.2) |
| Fully-adjusted (stratified by prior SUD) |  |
| *No prior SUD* | **4.7** (4.2 - 5.1) |
| *Prior SUD* | **2.3** (1.9 - 2.7) |
| Fully-adjusted (stratified by violent victimisation history) |  |
| *No prior violence victimisation* | **4.3** (3.9 - 4.7) |
| *Prior violence victimisation* | **2.9** (2.2 - 3.6) |

Minimally-adjusted: implicitly adjusted for age, biological sex, and county of residence in the year of diagnosis through matching. N missing individuals on parental characteristics: 372

**Table S6:** Only primary diagnosis of PTSD

| **Model specification** | **Matched cohort**  **(N = 76,417)** |
| --- | --- |
|  | HR (95% CI) |
| Minimally-adjusted | **7.8** (6.9 - 8.7) |
| Parental characteristics | **7.6** (6.7 - 8.5) |
| Parental characteristics + psychiatric comorbidities | **4.9** (4.3 - 5.7) |
| Parental characteristics + psychiatric comorbidities + history of violent victimization (fully-adjusted) | **4.4** (3.8 - 5.1) |
| Fully-adjusted (stratified by sex) |  |
| *Male* | **3.3** (2.6 - 4.3) |
| *Female* | **5.0** (4.2 - 5.8) |
| Fully-adjusted (stratified by prior SUD) |  |
| *No prior SUD* | **5.1** (4.3 - 5.9) |
| *Prior SUD* | **2.2** (1.5 - 3.1) |
| Fully-adjusted (stratified by violent victimisation history) |  |
| *No prior violence victimisation* | **4.6** (3.9 - 5.4) |
| *Prior violence victimisation* | **3.2** (2.0 - 5.0) |

N missing individuals on parental characteristics: 110

**Figure S1**: Derivation of matched cohort

**Individuals born in Sweden from 1973 to 2005**

n = 3,372,126

**Individuals eligible for inclusion in study**

n = 3,317,819

**Matched cohort**

**Exposed** (PTSD diagnosis): n = 29,688

**Unexposed** (no PTSD diagnosis): n = 296,880

*Excluded from study (n = 54,307):*

Adopted individuals (n = 12,521)

Information missing from at least one parent (n = 41,786)
